# Genetic structure of SARS-CoV-2 in Western Germany reflects clonal superspreading and multiple independent introduction events

**DOI:** 10.1101/2020.04.25.20079517

**Authors:** Andreas Walker, Torsten Houwaart, Tobias Wienemann, Malte Kohns Vasconcelos, Daniel Strelow, Tina Senff, Lisanna Hülse, Ortwin Adams, Marcel Andree, Sandra Hauka, Torsten Feldt, Björn-Erik Jensen, Verena Keitel, Detlef Kindgen-Milles, Jörg Timm, Klaus Pfeffer, Alexander T Dilthey

## Abstract

We whole-genome sequenced 55 SARS-CoV-2 isolates from Western Germany and investigated the genetic structure of SARS-CoV-2 outbreaks in the Heinsberg district and Düsseldorf. While the genetic structure of the Heinsberg outbreak indicates a clonal origin, reflective of superspreading dynamics during the carnival season, distinct viral strains are circulating in Düsseldorf, reflecting the city’s international links. Limited detection of Heinsberg strains in the Düsseldorf area despite geographical proximity may reflect efficient containment and contact tracing efforts.

---

Since its emergence in the Chinese city of Wuhan in late 2019, severe acute respiratory coronavirus 2 (SARS-CoV-2) has infected more than 2 million individuals and led to more than 130,000 deaths worldwide^1^. More than 10,000 globally sourced SARS-CoV-2 genomes are publicly available, and powerful data sharing and analysis platforms like GISAID^2^ and Nextstrain^3^ enable the collaborative analysis of viral population structure on a global level. Additional insights into transmission dynamics can be gained from focused investigations of individual outbreaks and by integrating genomic data with classical epidemiology.

Here we report on the genetic structure of SARS-CoV-2 in North-Rhine Westphalia, Germany’s most populous state. Our analysis includes the “Heinsberg outbreak” – comprising a superspreading event at a carnival session in Gangelt, a small municipality of about 12,000 inhabitants on the border between Germany and the Netherlands – and subsequent outbreak dynamics in the state capital Düsseldorf, located 70km from Gangelt and an international economic and air travel hub of about 600,000 inhabitants.

## SARS-CoV-2 Genome Sequencing

The Institute of Virology at Düsseldorf University Hospital was one of the first labs to offer SARS-CoV-2 diagnostics in Western Germany. 55 SARS-CoV-2 isolate samples, 10 directly linked to the Heinsberg outbreak (obtained from medical practices in the Heinsberg district or from residents of Heinsberg district patients treated at Düsseldorf University Hospital) and 45 from the city of Düsseldorf and surrounding districts, were acquired from diagnostic swabs sent to the Institute of Virology at Düsseldorf University Hospital. RNA extraction and reverse transcription were carried out as previously described^4^. DNA amplification and sequencing on the Oxford Nanopore platform were carried out according to the Artic protocol^5,6^ (Supplementary Text), yielding between 31 and 582Mb of raw sequencing data per sample (Supplementary Table S1). Bioinformatic analysis was based on the Artic pipelines and additional manual curation was carried out (Supplementary Text), yielding completely resolved genomes with 2 – 13 variant positions (Supplementary Table S2) relative to the SARS-CoV-2 reference genome^7^. Of note, we observed evidence for multi-allelic variant positions in 11 of 55 samples (Supplementary Table S2); for one such sample (NRW-39; 13 positions called as multi-allelic), PCR was repeated and a separate sequencing run was carried out, confirming the detected multi-allelic variant positions (Supplementary Text). Further work is necessary to investigate whether multi-allelic variant calls represent true within-patient strain variation. In a proof-of-concept experiment, we also successfully sequenced reverse-transcribed viral cDNA from patient material without an intermediate PCR-based amplification step (Supplementary Text), potentially enabling simplified sample preparation and increased read lengths for some samples in the future. Our study was IRB-approved by the ethics committee of the Heinrich Heine University Düsseldorf (#2020-839).

### Analysis of the Heinsberg Outbreak

First cases of SARS-CoV-2 infection in Germany were detected in late January 2020 and could be linked to recent travel to Northern Italy and China^8^. On 24^th^ and 25^th^ February 2020, however, a married couple from the Heinsberg district with no known travel history to SARS-CoV-2 risk areas were diagnosed with SARS-CoV-2; by 28^th^ February 2020, the number of confirmed infections in the Heinsberg district had grown to 37; by 22^th^ April 2020, to >1,700^9^. Contact tracing later showed that many of the early SARS-CoV-2 cases could be linked to a carnival session in the municipality of Gangelt, part of the Heinsberg district, held on 15^th^ February 2020^8^. The “Heinsberg outbreak” represented one of the first large-scale SARS-CoV-2 outbreaks in Germany, seeded by community transmission and amplified by superspreading-type dynamics. Genomic analysis of 10 SARS-CoV-2 isolates from the Heinsberg outbreak, sampled between 25^th^ and 28^th^ February and including the first two diagnosed cases, demonstrated the clonal origin of the outbreak (Figure 1); all Heinsberg samples shared the same 2 stem mutations (Supplementary Table S2). Viral diversity in the Heinsberg samples varied between 2 and 6 variants relative to the SARS-CoV-2 reference genome^7^, and 5 distinct viral haplotypes could be identified (Supplementary Table S2). An analysis (Supplementary Text) of other publicly available SARS-CoV-2 sequences did not reveal an obvious origin of the Heinsberg outbreak (Supplementary Table S4); the Heinsberg isolates are not related to early sequences from other German outbreak areas (Bavaria, Baden Wuerttemberg), and, despite intense Dutch viral sampling efforts (585 available viral genomes from the Netherlands at the time of analysis), our analysis identified only 2 closely related isolates from the Netherlands (one collected on 21^st^ March, the other with undefined collection date). The role of the first two patients’ short vacation in the Netherlands seven days prior to the Gangelt carnival session^10^ thus remains, while suggestive in terms of reported SARS-CoV-2 incubation periods^11^, ambiguous. What is more, large numbers of closely related isolates are circulating in many countries, for example England, Wales, and Iceland (Supplementary Table S4). The small number of stem variants (2) compared to the maximum number of per-isolate variants (6) in the Heinsberg isolates, likely acquired over a period of a few weeks, is compatible with a relatively recent introduction from China.

**Figure 1.**
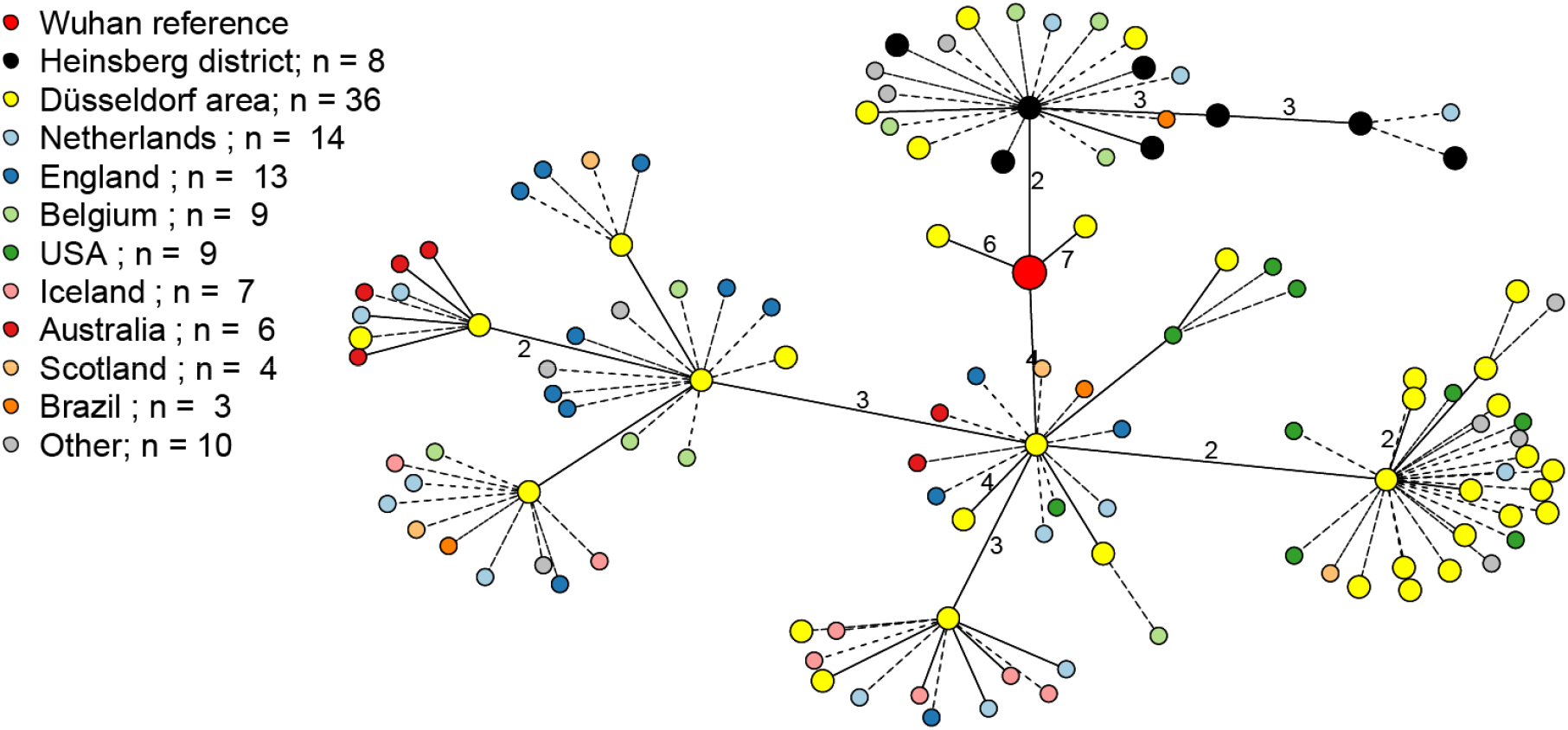
Minimum spanning tree showing 44 unambiguously resolved (see below) SARS-CoV-2 genomes from the Heinsberg district (n = 8) and the Düsseldorf area (n = 36); the original Wuhan SARS-CoV-2 reference genome; and a sample of closely related publicly available SARS-CoV-2 samples from GISAID (distance to any of the Heinsberg/Düsseldorf genomes of 0 or 1; see Supplementary Text for details). 11 isolate genomes from our study with multi-allelic variant calls are not considered “unambiguously resolved” for the purpose of this analysis and thus omitted. Dashed and solid edges without adjacent numbers indicate distances of 0 and 1, respectively; all other distances are shown explicitly.

### Düsseldorf Outbreak Dynamics

The first SARS-CoV-2 cases in Düsseldorf, 70km from Gangelt, were diagnosed in early March 2020^12^; as of 21^st^ April 2020, the outbreak had grown to more than 900 cases^13^. The set of 55 whole-genome-sequenced isolates included 45 samples from Düsseldorf and nearby districts, collected between 3^rd^ and 23^rd^ March. A minimum spanning tree analysis of unambiguously resolved viral sequences (Figure 1) showed that there were at least 5 clusters of viral haplotypes circulating in the Düsseldorf area; the number of variant positions relative to the SARS-CoV-2 reference genome in the Düsseldorf samples varied between 2 and 13 (Supplementary Table 2). Closely related strains (distance 0 or 1) were found in the United States, the United Kingdom, Australia, and many other countries (Supplementary Table S4), strongly indicating multiple independent introduction events. Of note, 4 “Düsseldorf area” isolates clustered with the Heinsberg outbreak (Figure 1); of these, 2 were collected from residents of a district next to Heinsberg treated at Düsseldorf University Hospital, and 2 remained of unclear origin (patient data not available). Thus, there was no evidence for widespread community circulation of Heinsberg-derived SARS-CoV-2 strains in the Düsseldorf area.

### Illumina Validation

To verify the accuracy of Nanopore-based viral assembly, additional Illumina sequencing was carried out for the first 11 samples of our cohort (Supplementary Table S1; Supplementary Text); data analysis was carried out with iVar^14^. For 41 of 45 variant positions identified by either Nanopore or Illumina, the called variant alleles agreed; manual inspection of the discordant positions revealed low coverage for 2 discordant positions and one missed multi-allelic call for each approach (Supplementary Table S3).

## Discussion

As SARS-CoV-2 case numbers and the social and economic consequences of social distancing and lock-down measures continue to rise, many countries are facing difficult trade-offs. Improved methods to characterize the dynamics of viral transmission are urgently required. Here we have investigated the genetic structure of two related SARS-CoV-2 outbreaks in Western Germany using Nanopore sequencing, which has additional applications in many fields such as human genetics^15^ and microbial metagenomics^16^. We have demonstrated the clonal origin of the Heinsberg outbreak, consistent with existing epidemiological data on a carnival session in Gangelt as the epicentre of the outbreak. The lack of association between the Heinsberg samples and other early German outbreak isolates is suggestive of a separate introduction event, possibly via the Netherlands, China, or a third country. By contrast, SARS-CoV-2 isolates circulating in Düsseldorf are highly polyclonal and can be grouped into at least 5 clusters of viral haplotypes. Despite the geographical proximity between Heinsberg and Düsseldorf, only 4 of 36 unambiguously resolved samples from the Düsseldorf area clustered with the Heinsberg outbreak, and 2 of these were derived from residents of a district neighbouring Heinsberg. Limited detection of Heinsberg strains in the Düsseldorf area may reflect the efficacy of the contact tracing efforts conducted by the German public health authorities; of note, “lockdown”-type restrictions with limits on public gatherings in Germany were only imposed on 23^rd^ March 2020^17^, i.e. on the day on which the last sample of our study was collected. More extensive sampling of SARS-CoV-2 isolates from Western Germany will be required to investigate the effect of various containment measures on transmission chains at a genomic level. Consistent with reports from Iceland^18^, New York^19^, and data on Nextstrain, our study has demonstrated the simultaneous circulation of distinct viral haplotypes in a metropolitan region; even within the Heinsberg outbreak, we could identify 5 distinct viral haplotypes. Importantly, as SARS-CoV-2 genomes continue to diverge as part of ongoing viral evolution, the application of genomic epidemiology^20,21^ for the identification and targeted interruption of viral transmission chains will become increasingly feasible.

## Data Availability

All generated viral genome assemblies have been submitted to GISAID; all generated assemblies and the raw sequencing data are also available on NCBI (BioProject PRJNA627229).

## Acknowledgements and Data Availability

This work was supported by the Jürgen Manchot Foundation and by funding from the German Federal Ministry of Education and Research (Bundesministerium für Bildung und Forschung; Award number 031L0184B). We gratefully acknowledge the Authors, the Originating and Submitting Laboratories for their sequence and metadata shared through GISAID. All submitters of data may be contacted directly via <u>GISAID</u>. The Acknowledgments Table for GISAID is part of the Supplement (Supplementary Table S5). We would also like to thank Nicholas Loman and Josh Quick for advice and discussions.

## Supplementary Table Legends

**Supplementary Table S1: Sample and sequencing data summary**. The table shows sequencing data statistics for each sample and sequencing technology; RT-PCR Ct values, quantifying viral load in the diagnostic sample that was used for RNA extraction; and NCBI accession IDs. The IDs specified in the column “Sequencing Run” correspond to identifiers submitted to GISAID.

**Supplementary Table S2: Assembly and variant summary**. The table shows the detected variants in each sample as well as sampling date, an assigned location, and assembly summary statistics, such as the number of undefined (“N”) characters in each assembly. In the variant table, multiallelic variant positions are highlighted with a lighter gray. Samples designated as “Heinsberg district” were sent in from medical practices in the Heinsberg district or obtained from residents of Heinsberg district treated at Düsseldorf University Hospital; all other samples are from the Düsseldorf area.

**Supplementary Table S3: Illumina / Nanopore comparison**. The table shows, for the 11 samples for which both Nanopore and Illumina sequencing were carried out, the positions and called genotypes at all variant positions. For the purposes of this table, “variant positions” are defined as positions at which iVar detects a variant or at which the Nanopore-based assembly submitted to GenBank/GISAID contains a non-N non-reference allele. “Reported”: Allele present in the Nanopore-based assembly submitted to GenBank/GISAID, after manual curation; “Nanopolish”: allele called by Nanopolish; “Medaka”: allele called by Medaka; “iVar”: allele called by iVar, i.e. based on the Illumina data. Positions with discordant calls were manually inspected.

**Supplementary Table S4: Genomic neighbourhood of Düsseldorf/Heinsberg samples**. The table shows, for each unambiguously resolved Düsseldorf/Heinsberg sample (i.e., for each sample with no other ambiguous nucleotide characters than “N”), the full set of determined genomic neighbours, defined as the set of unambiguously resolved non-Düsseldorf/Heinsberg sequences from GISAID with distance 0 or 1, and aggregated country-of-origin statistics for these sequences. See Supplementary Text for details.

**Supplementary Table S5: Acknowledgement table for GISAID**.

## Notes

### Competing Interest Statement

The authors have declared no competing interest.

## References

1. World Health Organization. Coronavirus disease 2019 (COVID-19) Situation Report – 88. https://www.who.int/docs/default-source/coronaviruse/situation-reports/20200417-sitrep-88-covid-191b6cccd94f8b4f219377bff55719a6ed.pdf?sfvrsn=ebe78315_6 (2020)

2. Shu, Y. & McCauley, J. GISAID: Global initiative on sharing all influenza data - from vision to reality. Euro Surveill 22(2017).

3. Hadfield, J. et al. Nextstrain: real-time tracking of pathogen evolution. Bioinformatics 34, 4121–4123 (2018).

4. Walker, A., Ennker, K.S., Kaiser, R., Lubke, N. & Timm, J. A pan-genotypic Hepatitis C Virus NS5A amplification method for reliable genotyping and resistance testing. J Clin Virol 113, 8–13 (2019).

5. Quick, J. ARTIC amplicon sequencing protocol for MinION for nCoV-2019. https://dx.doi.org/10.17504/protocols.io.bbmuik6w (2020)

6. Quick, J. et al. Multiplex PCR method for MinION and Illumina sequencing of Zika and other virus genomes directly from clinical samples. Nat Protoc 12, 1261–1276 (2017).

7. Wu, F. et al. A new coronavirus associated with human respiratory disease in China. Nature 579, 265–269 (2020).

8. Robert Koch Institute. Coronavirus Disease 2019 (COVID-19) Daily Situation Report 04/03/2020. https://www.rki.de/DE/Content/InfAZ/N/Neuartiges_Coronavirus/Situationsberichte/2020-03-05-en.pdf?blob=publicationFile (2020)

9. Kreisverwaltung Heinsberg. Coronavirus im Kreis Heinsberg. https://www.kreis-heinsberg.de/aktuelles/aktuelles/?pid=5149 (2020)

10. Netherlands National Institute for Public Health and the Environment. Duitse coronapatiënt niet ziek tijdens verblijf in Limburg. https://www.rivm.nl/nieuws/duitse-coronapatient-niet-ziek-tijdens-verblijf-in-limburg (2020)

11. Lauer, S.A. et al. The Incubation Period of Coronavirus Disease 2019 (COVID-19) From Publicly Reported Confirmed Cases: Estimation and Application. Ann Intern Med (2020).

12. Pressedienst Landeshauptstadt Düsseldorf. Zwei Düsseldorfer mit Coronavirus infiziert. https://www.duesseldorf.de/medienportal/pressedienst-einzelansicht/?L=0 (2020)

13. Pressedienst Landeshauptstadt Düsseldorf. Die Coronazahlen vom 21. April. https://www.duesseldorf.de/medienportal/pressedienst-einzelansicht.html (2020)

14. Grubaugh, N.D. et al. An amplicon-based sequencing framework for accurately measuring intrahost virus diversity using PrimalSeq and iVar. Genome Biol 20, 8 (2019).

15. Jain, M. et al. Nanopore sequencing and assembly of a human genome with ultra-long reads. Nat Biotechnol 36, 338–345 (2018).

16. Dilthey, A.T., Jain, C., Koren, S. & Phillippy, A.M. Strain-level metagenomic assignment and compositional estimation for long reads with MetaMaps. Nat Commun 10, 3066 (2019).

17. Robert Koch Institute. Coronavirus Disease 2019 (COVID-19) Daily Situation Report 23/03/2020. https://www.rki.de/DE/Content/InfAZ/N/Neuartiges_Coronavirus/Situationsberichte/2020-03-23-en.pdf?blob=publicationFile (2020)

18. Gudbjartsson, D.F. et al. Spread of SARS-CoV-2 in the Icelandic Population. N Engl J Med (2020).

19. Gonzalez-Reiche, A.S. et al. Introductions and early spread of SARS-CoV-2 in the New York City area. medRxiv, 2020.04.08.20056929 (2020).

20. Grubaugh, N.D. et al. Tracking virus outbreaks in the twenty-first century. Nat Microbiol 4, 10–19 (2019).

21. Gardy, J.L. & Loman, N.J. Towards a genomics-informed, real-time, global pathogen surveillance system. Nat Rev Genet 19, 9–20 (2018).

